# Study protocol for microneurographic investigation of nociceptor sensitisation in Fibromyalgia Syndrome. (MICRO-FMS)

**DOI:** 10.64898/2026.02.24.26346973

**Authors:** Elise A. Ajay, Faris Khan, Atanu Bhattacharjee, Anthony E. Pickering, James P. Dunham

**Affiliations:** Anaesthesia, Pain, and Critical Care Sciences, School of Physiology, Pharmacology, & Neuroscience, University of Bristol; Population Health and Genomics, School of Medicine, University of Dundee, Dundee, UK

## Abstract

**Introduction:** Chronic pain in fibromyalgia may be driven by abnormal ongoing activity in a subclass of C-fibre nociceptors known as Type1B or CMi nociceptors. As is common in C-nociceptor microneurography studies, the modest patient numbers in these prior studies generate large confidence intervals around the point estimate of the prevalence of this abnormal activity. This complicates the interpretation of the relative importance of this ongoing nociceptor activity as a pain generating mechanism in fibromyalgia. The study aims to improve precision via an adaptive Bayesian protocol that maximises the yield and quality of data collection whilst minimising patient burden.

**Methods:** The study employs an optimised microneurography protocol with an adaptive study design. The microneurography protocol incorporates early identification of CMi nociceptors via an abbreviated activity dependent slowing protocol to increase yields enabling efficient collection of the primary outcome data. The adaptive study design will use Bayesian principles to iteratively assess the predictive probability of futility, and terminate early if there is high confidence that the hypothesis is false. Furthermore, the study will employ questionnaires to explore links with pain in the area under study to the electrophysiology data. Finally, quantitative sensory testing will be used to investigate whether the irritable nociceptor phenotype is associated with abnormalities in CMi nociceptor physiology.

**Ethics & Dissemination:** This study has received HRA REC approval in the UK. Participants will provide written informed consent, and may withdraw at any time without consequence. At the end of the study, the results will be disseminated through peer-reviewed publication, and the data made available via a data repository.

**Strengths & limitations of this study:**

Bayesian predictive probability of futility to minimise patient burden in microneurography
Microneurography for objective interrogation of the peripheral nervous system
Optimised microneurography protocol to efficiently answer primary hypotheses
Subjective elements of early termination criteria of the study assessed and co-developed with Patient and Public Inclusion and Engagement Group

## Introduction

Fibromyalgia is a primary chronic pain syndrome characterised by widespread pain, persisting for more than 3 months (Clauw, 2024; Nicholas et al., 2019). The prevalence is estimated to be between 2 and 8% (Fayaz et al., 2016; Heidari et al., 2017; Walitt et al., 2015). It is associated with significant emotional distress and functional disability with symptoms including fatigue, cognitive dysfunction, and sleep disturbance (Berwick et al., 2022).

The aetiology of fibromyalgia remains poorly understood (Arnold et al., 2016). The condition may arise from pathophysiological changes in either, or both, the central and peripheral nervous systems (Nijs et al., 2021; Siracusa et al., 2021). Indeed, there are consistent reports of periphery abnormalities in patients with fibromyalgia including histological, quantitative sensory testing findings, and clinical questionnaires indicative of small fibre pathology, (Grayston et al., 2019; Kim et al., 2008; Marshall et al., 2024; Üçeyler et al., 2013). More recently, altered tactile afferent function has also been reported (Israel et al., 2025).

Critically, microneurography studies have reported hyperexcitability in peripheral C-fibre nociceptors in patients with fibromyalgia (Evdokimov et al., 2019; Serra et al., 2014). Specifically, 40.7% (n = 11/27) of patients were shown to have abnormal spontaneous activity in Type 1B (also referred to as mechanically-insensitive C-fibre (CMi) nociceptors), but not in other C-nociceptors. Although spontaneous activity in C-fibres is a normal feature of aging, it is typically only seen in 2-7.7% of healthy adults (Evdokimov et al., 2019; Namer et al., 2009; Ribeiro et al., 2025; Serra et al., 2014). In addition, these studies also show that Type 1B fibres develop abnormal mechanical sensitivity in 19% of fibromyalgia patients, compared with 7% of healthy age matched controls.

It has therefore been proposed that sensitisation and spontaneous activity in Type 1B nociceptors in patients with fibromyalgia represents the primary pain generator (Evdokimov et al., 2019; Serra et al., 2014). For comparison, in patients with painful polyneuropathy (i.e., neuropathic pain), spontaneous activity with or without mechanical sensitisation, was seen in 25% of CMi fibres and 64% of patients (n = 7/11) whereas in patients with non-painful polyneuropathy (n = 8) these changes were seen in 12% of CMi fibres and 50% of patients (Kleggetveit et al., 2012).

In microneurography, C-fibre nociceptors and their subtypes can be distinguished from other afferent and efferent C-fibres using activity dependent slowing (ADS) (for review see (Namer & Lampert, 2025)). This is very useful in the context of low signal:noise ratios that are typical in C-fibre recordings. ADS refers to the phenomenon that repeated activation of a peripheral nerve fibre decreases their conduction velocity. Multiple studies have established that C-fibre afferents, including nociceptors, can be identified and distinguished using electrical stimulation alone in healthy participants (Obreja et al., 2010; Serra et al., 1999; Watkins et al., 2017), and that this method of categorisation is resilient even in disease (Namer et al., 2015; Ørstavik et al., 2006; Serra et al., 2012).

Type 1B and CMi are terms used to describe C-fibres with large degrees of ADS at both low and high frequencies, that conduct in the C-fibre latency range. They are normally mechanically insensitive, except for prior sensitisation by inflammation or other agents. For simplicity, we will henceforth refer to these fibres as CMi fibres.

ADS can also be utilised to identify non-electrically evoked activity. For example, on a background of low frequency electrical stimulation, mechanical stimuli sufficient to excite the nociceptor manifests as a slowing of responses to subsequent electrical stimuli. This is known as the marking technique (Torebjörk & Hallin, 1974). In pathology, this technique can be used to identify abnormal responses to mechanical stimulation, such as identification of sensitised CMi fibres as described above.

Furthermore, if the nociceptor has spontaneous or ongoing activity, this may manifest as a ‘jitter’ in the constant latency response, i.e., the electrically evoked action potentials are slowed to differing degrees depending on the frequency of preceding non-electrically evoked activity (Serra et al., 2014). ADS has therefore been critically important for the identification of both pathological sensitisation and spontaneous activity in C-nociceptors in fibromyalgia (Evdokimov et al., 2019; Serra et al., 2014).

Sensory afferent function can also be assessed psychophysically using quantitative sensory testing (QST). This approach has been applied to multiple chronic pain pathologies for treatment stratification (Demant et al., 2014), including for fibromyalgia (Marshall et al., 2024; Rehm et al., 2021). Abnormal QST measures in a patient population alone does not necessarily indicate the mechanism underlying chronic pain (Forstenpointner et al., 2021), however the approach does provide important quantification of loss- and gain-of function in somato-sensory perception (Backonja et al., 2013).

Microneurography has traditionally been considered a high quality, but low data yield, research approach. This view is now being challenged (Serra, 2010), with microneurography-guided clinical trials (Serra et al., 2015), reference datasets (Ribeiro et al., 2025), and with recent comprehensive reviews (Namer & Lampert, 2025).

However, it does remain time consuming and can be demanding for both researcher and participant (Vallbo, 2018). This means that replication studies in microneurography are rare, risk being under powered, and the original studies themselves often have modest participant numbers.

Bayesian study designs enable the iterative incorporation of new data with prior evidence to update the estimated probability of a predicted outcome (Berry, 2025; Bhattacharjee, 2020; Lee, 2024). Such approaches also allow for interim assessments of efficacy or futility, thereby improving research efficiency. This is not only through optimising cost and investigator time, but also by optimising participant involvement through minimisation of unnecessary exposure, risk, and burden. This is relevant as though microneurography has been shown to be a safe technique, adverse events can occur. For example it is estimated that 4.6% and 2.8% of participants experience paraesthesia and pain respectively after microneurography and that this can last up to several weeks (Meah et al., 2019).

## Methods & Analysis

This study aims to test for the presence of abnormal spontaneous activity and mechanical excitability in C-mechano-insensitive (CMi) nociceptors in patients with fibromyalgia. Furthermore, the study will use questionnaires and quantitative sensory testing (QST) to examine the relationship between the patient’s pain report, sensory function, and electrophysiology. The protocol will apply a Bayesian approach to stop recruitment early in the event of statistical futility.

### Ethics and informed consent

The patient study has been approved by the South Central - Hampshire B Research Ethics Committee on behalf of the Health Research Authority (REC reference number: 24/SC/0386, Protocol number 2024-5219). The healthy volunteer study has been approved by the Faculty of Life Science and Science Research Ethics Committee, University of Bristol (Ref 51882). Other than recruitment route, the patient and healthy volunteer protocols are identical. All participants will provide written informed consent and will be reimbursed for their time and travel costs. The study is sponsored by the University of Bristol, UK.

### Participant recruitment

Patients will be recruited (via posters, clinician referral, and database searches) from secondary care Pain Clinics at North Bristol NHS Trust and at University Hospitals Bristol and Weston NHS Foundation Trust starting in 2025. Upon receipt of initial interest from a patient, they will be provided with a participant information sheet and study leaflet. Age- and sex- matched controls will be recruited from the Bristol area through posters and online advertisements. All participants who volunteer to take part will undergo telephone screening for the following inclusion and exclusion criteria:

### Inclusion criteria

- Adults, >18 years of age
- Patient has fibromyalgia (for the patient arm of the study)
- Participant has capacity to consent
- Participant is able to communicate in English, as is necessary for the safe conduct of the intraneural recordings, and accurate completion of the patient questionnaires and sensory testing.

### Exclusion criteria

- Unable to tolerate microneurography, e.g., needle phobia, unable to lie still, history of stress/fear/pain-induced syncope
- Higher risk from microneurography, for example:

- Neurological disorders such as Multiple Sclerosis, Parkinson’s Disease
- Infection/oedema/broken skin over recording or stimulation site
- Cardiac or other pacemaker/stimulation device
- Taking anti-coagulants/altered coagulation
- Are deemed to be unsuitable for participation in the study in the opinion of the study investigators

Patients will then discuss the study further with the investigators, including any risks, and have the opportunity to ask questions. If they wish to continue, they will be invited to provide written informed consent.

## Data collection and management

All study data, excluding the raw microneurographic data recordings but including the microneurographic measures (see below), will be collected using REDCap (Research Electronic Data Capture) (Harris et al., 2009, 2019) and stored at the University of Bristol. REDCap is a secure, web-based data collection tool frequently used in clinical and translational research allowing data anonymisation, streamlined and validated data collection, audit trails, and automated export procedures.

### Questionnaires and tender point examination

Participants’ symptom burden will be assessed using questionnaires including: the Core Minimum Dataset (Laskawska et al., 2022), an adapted short-form McGill Pain Questionnaire, the Fibromyalgia Impact Questionnaire Revised (Bennett et al., 2009), the Royal College of Physicians Fibromyalgia Diagnostic Worksheet (Wolfe et al., 2016), Douleur Neuropathique 4 Questions (Bouhassira et al., 2005), and the Brief Pain Inventory (Cleeland, 1991). Questionnaires are simplified to minimise duplicate questions.

The short-form McGill Pain Questionnaire is adapted to specifically capture pain descriptors, current pain (via a numeric rating scale (NRS)), and perceived source of pain (Superficial i.e. the skin; Deeper structures i.e., muscles, bones, joints; Unsure; Not Applicable) *as experienced in the body area that will be tested in the microneurography*. Most often this will be the dorsum of the foot, via the superficial peroneal nerve, as per the prior studies with microneurography in fibromyalgia.

The location of the participants’ (widespread) pain will be captured electronically using the CHOIR body map (Scherrer et al., 2021). Current medications for pain and/or other conditions will also be recorded. Participants will be examined for tender point count as per the American College of Rheumatology 1990 criteria (Wolfe et al., 1990). The questionnaires and examination will take approximately 20 minutes to complete.

### Quantitative sensory testing

Participants will then be assessed using quantitative sensory testing (see protocol overview in Figure 1). Assessors will not be blinded to the participant’s patient or control status. The examination aspects of the Douleur Neuropathique 4, i.e., assessment for hypoaesthesia to touch and pin prick, and brush allodynia will be assessed within the QST session. Test instructions will be read to the patient, in English, to ensure consistency. Unless otherwise stated, QST stimulation will be applied to the site contralateral to the planned microneurography testing site – typically the right foot dorsum. The assessment of thermal thresholds, mechanical thresholds and mechanical pain sensitivity, are based upon the German Research Network on Neuropathic Pain (DFNS) protocol (Rolke 2006).

**Figure 1.**
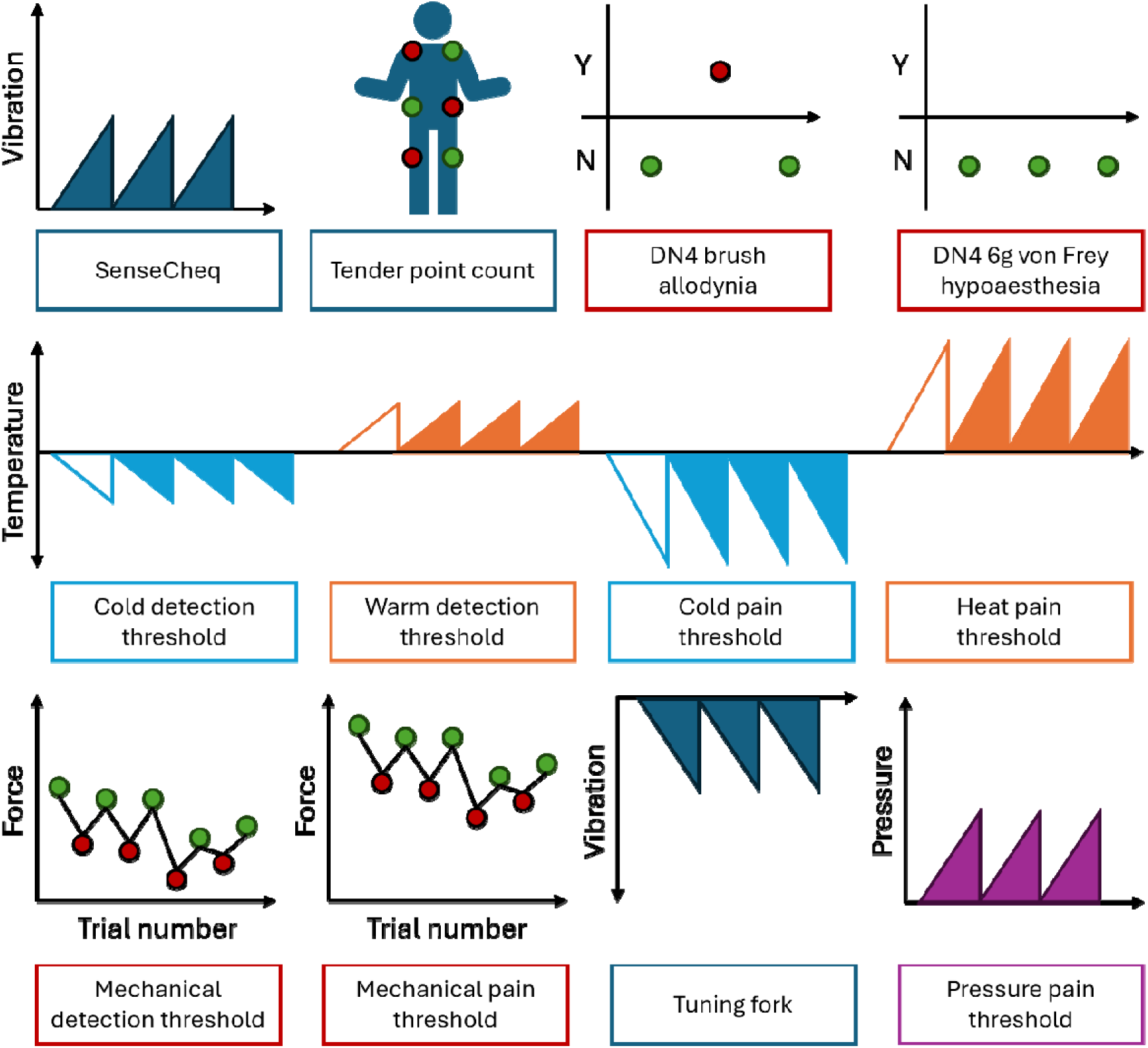
Examination and quantitative sensory testing protocol. See text for further details.

**Figure 2.**
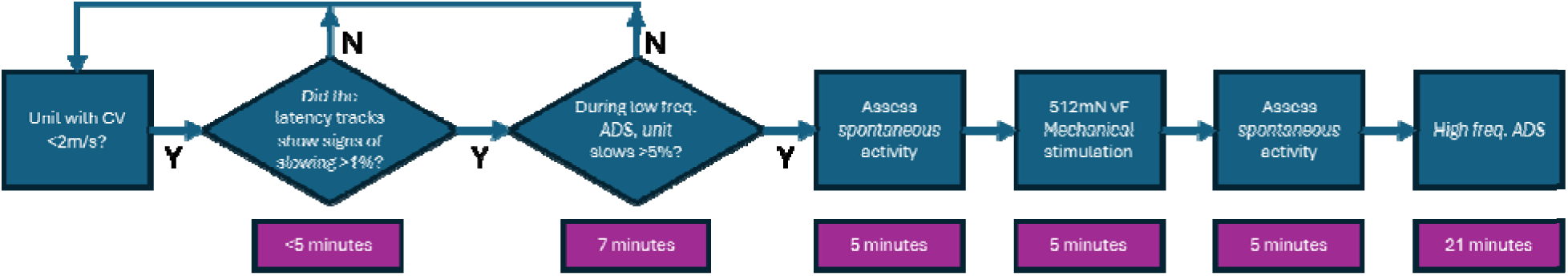
Microneurography protocol. Two checks in the early stages of the protocol will maximise yield o Type 1B/CMi fibres whilst minimising experiment duration.

Warm detection, cold detection, heat pain, and cold pain thresholds will be assessed using a thermal stimulator with a total area size of 9 cm^2^ (QST.Labs TCSII, probe T11) placed on the skin. Starting from a neutral temperature (32°C), the thermode temperature will ramp at 1°C per second until the participant reports either a detection of the temperature change (detection thresholds) or detection of pain (pain thresholds) via a button press. Following the button press, the thermode will return to neutral temperature, and the next trial will begin after a randomised period of 4-6 seconds for detection thresholds or 10 seconds for pain thresholds. Each test will be repeated 4 times, with the first ramp discarded as training.

Thresholds for innocuous mechanical stimuli will be assessed using calibrated von Frey filaments (Touch Test Sensory Probes, Stoelting Co. USA) using the method of limits. Mechanical pain thresholds, using the method of limits and the stimulus-response-functions (i.e., mechanical pain sensitivity), will be assessed using calibrated punctate stimulators (8-512 mN, PinPrick stimulators, MRC Systems, Germany). For mechanical pain sensitivity, stimulators will be presented in random order (generated in REDCap), and participants will rate the perceived pain on a 0-100 numerical rating scale, where 0 is “no pain” and 100 is “most intense pain imaginable”. Thresholds and sensitivity trials will be repeated 5 times each.

Brush allodynia will be assessed using three trials of gentle brushing while capturing the participants’ perception of any pain.

Pressure pain thresholds will be assessed using a pressure algometer (Somedic, Sweden) with a 1 cm^2^ contact area. The algometer will be applied three times to a large muscle group near, but contralateral to, the microneurography testing site (e.g., the tibialis anterior of the right leg), using slowly increasing force (50 kPa/s).

Vibration perception threshold test will be performed using a Rydel-Seiffer tuning fork applied to the contralateral testing site. A second vibration detection threshold measure will also be obtained for the fingertip using a custom-built device, ‘SenseCheQ’ (Dujmović et al., 2025), that delivers ascending vibratory stimuli via a haptic actuator.

The participant is instructed to push a button with the opposite hand on detection of the vibration.

The complete set of quantitative sensory testing procedures will take approximately 45 mins to complete.

### Microneurography

Microneurography will be performed as previously described (Dunham et al., 2018). Electrode pairs with an active electrode impedance of 2 MΩ and 200 µm diameter will be used (Frederick Haer, Bowdoin, ME, USA. Cat No. UNP35F2U). The active electrode will be inserted into the superficial peroneal nerve proximal to the lateral malleolus under ultrasound guidance. The reference electrode will be inserted subcutaneously and in close proximity to the recording electrode. The patient’s wellbeing will be monitored throughout the microneurography procedure, particularly for signs of vasovagal syncope (e.g., paleness, sweating, etc.), by a team member who is not performing microneurography. If such signs or symptoms occur the electrodes will be removed and the participant laid flat in the bed until they recover. The experiment will then cease. Acute and delayed side effects will be actively sought via questionnaire immediately after the experiment and 1-2 weeks later for all participants.

Differential recordings will be made, amplified, and digitised, using an Intan RHD 2116 chip sampled at 30 kHz and passed to an OpenEphys acquisition system (http://open-ephys.org) and bandpass filtered (300-6000 Hz). The signal will be visualised using the OpenEphys graphical user interface. The data acquisition system is battery powered and the participant will be electrically isolated from the recording equipment via a USB 3.0 optoisolator (5 kV RMS isolation - <u>7055 Series USB Isolator (intona.eu)</u>).

The innervation area on the dorsum of the foot will be searched with electrical stimulation (square pulse, 0.5 ms duration, 0.25 Hz, 0-20 mA - intensity titrated to a participant pain score of ∼4/10 on the numeric rating scale) using needle electrodes (Ambu Neuroline twisted pair subdermal EEG electrodes, Denmark). When a slowly conducting unit with a stable latency is identified, the stimulating electrodes will be rested on the foot such that the tips are intradermal and do not move. The search for the innervation area and acquisition of single unit activity typically occurs within 20 minutes, and will not exceed an hour to minimise risk of persistent abnormal sensation (Eckberg et al., 1989). Latency raster plots will be visualised using the OpenEphys plug-in APTrack (Nickerson et al., 2023).

CMi fibres will be sought, and their characteristics explored using a combination of activity dependent slowing and mechanical stimulation via the marking technique. Fibres with stable long latency responses with conduction speeds consistent with C fibres (<2 m/s) will be identified. If these slow (i.e., the latency increases) by >1% during 0.25 Hz stimulation (Serra et al., 2004) they will be taken forward for further characterisation. If they do not, they will be discarded and the electrode position altered, and the skin searched again until a putative CMi fibre is identified.

When a putative CMi is identified, it will then be electrically stimulated at 2x electrical threshold (as tolerated by the participant), with a low frequency activity-dependent slowing protocol (Weidner et al 1999., Obreja et al., 2010), consisting of: 2 minutes rest (i.e., no electrical stimulation), 20 pulses at 0.125 Hz, 20 pulses at 0.25 Hz, 30 pulses at 0.5 Hz, 20 pulses at 0.25 Hz. If the fibre has slowed more than 5% at the end of the 0.5 Hz stimulation, with a recovery to baseline latency after the 0.5 Hz stimulation >44% within 10 electrical pulses (Beisswanger et al., 2022), in combination with the >1% slowing seen during initial characterisation, then the unit will be classified as a CMi. If the unit is not a CMi, it will be discarded, and a new putative CMi will be sought.

Spontaneous activity in CMi units will then be assessed. The electrical receptive field will be stimulated at 0.25 Hz, 2x electrical threshold for 5 minutes. If the constant latency tracks are disturbed by sudden or rounded perturbations >1.5% of the baseline latency they will be deemed to have additional spontaneous firing (Serra et al., 2012).

The primary outcome measure is the patient-level prevalence of spontaneous activity in CMi fibres among fibromyalgia patients. A patient will be classified as positive for spontaneous activity if at least one confirmed CMi fibre demonstrates spontaneous or ongoing activity according to the predefined electrophysiological criteria. The statistical unit of analysis is therefore the patient.

The unit will then be assessed for mechanical sensitivity. During 0.25 Hz stimulation at 2x electrical threshold, the skin area around the stimulation site (∼2 cm^2^; as tolerated) will be probed using a 512 mN von Frey filament (Optihair, MRC systems). During this mechanical stimulation the participant will be asked to quantify any mechanically evoked pain using an electronic visual analogue scale with “no pain” equal to zero and “worst imaginable pain” as equal to 10.

A further 5-minute period of 0.25 Hz electrical stimulation will follow mechanical stimulation. This period will be used to allow the identification of prolonged, non-electrically evoked activity following mechanical stimulation, as per the classification above of abrupt or rounded perturbations exceeding 1.5% of the constant latency responses, and as reported previously (Evdokimov et al., 2019).

Finally, the unit will be characterised using a high frequency activity dependent slowing protocol at 2x electrical threshold (as tolerated) consisting of: 2 minutes rest, 3 minutes at 0.25 Hz, 3 minutes rest, 6 minutes at 0.25 Hz, 3 minutes at 2 Hz, 6 minutes at 0.25 Hz. C-fibre nociceptors will slow >10% during the high frequency activity dependent slowing protocol (Serra et al., 1999).

At the end of the experiment, the conduction distance between the recording electrode and the receptive field of the single unit will be approximated using a measuring tape flattened against the skin. Patient wellbeing will be continuously monitored as described above and the patients will be actively followed up 1-2 weeks later.

### Recording analysis

Following the above in-experiment analysis, the acquired electrophysiological recordings will be processed and analysed offline. Spikes of single unit tracks will be identified using SpikeSpy (https://github.com/Microneurography/SpikeSpy). Using the spikes times of single unit tracks, the conduction properties of the unit (to confirm classification) will be calculated, including:

- Conduction velocity (CV)
- Percentage slowing and recovery during the low frequency ADS protocol
- Percentage slowing and recovery during the high frequency ADS protocol

In addition, we will assess for:

- The presence of spontaneous activity (primary outcome measure) will be defined by the observation of abrupt or rounded changes in latency exceeding 1.5% during the 0.25 Hz stimulation immediately following the low frequency ADS protocol, immediately following the mechanical stimulation, and at any time during the 0.25Hz stimulation periods of the high frequency protocol.
- Presence of responses to mechanical stimulation, i.e., abrupt changes in latency coinciding with mechanical stimulation followed by a gradual return to baseline latency (Schmelz et al., 1995).

These microneurographic measures will then be inputted into the REDCap database.

Patients in whom no CMi fibre can be identified despite completion of the microneurography protocol will be excluded from the primary analysis but reported descriptively. A sensitivity analysis will additionally classify such patients as not exhibiting spontaneous CMi activity to assess robustness of the primary findings.

### Statistics and interim analysis strategy

Interim analysis using a Bayesian approach will be used to determine if the study should be terminated early according to the probability of detecting a meaningful prevalence of spontaneous activity in fibromyalgia patients. This approach minimises burden on participants and researchers whilst balancing the risk of a false negative outcome.

To explore a meaningful prevalence of spontaneous activity in patient with fibromyalgia, we reviewed the published literature on spontaneous activity in C-fibres in healthy aged populations, and in patients. In healthy aged populations, spontaneous activity is reported to be present in the Type1B/CMi fibres in:

- 2.2% in <u>participants</u> aged 40 ± 4 years (1/45, 95% CI: 0-12%; Clopper-Pearson interval) (Serra et al., 2014)
- 7.7% <u>participants</u> (1/13, 95% CI: 0-36%; Clopper-Pearson interval) of median age 42 (range 24-61 years) (Evdokimov et al., 2019)
- in 5 of 142 <u>afferent C-fibres</u> from 15 participants with a mean age 56 years (range 41–67 years) (Namer et al., 2009).

These publications suggest that 2.2-7.7% of healthy aged people have spontaneous activity in Type 1B/CMi fibres, but note that the confidence intervals around this estimate are high.

In contrast, in patients with fibromyalgia, spontaneously active Type 1B/CMi fibres were found in 11 of 27 patients (40.7%, 95% CI: 22-61%; Clopper-Pearson interval) (Evdokimov et al., 2019).

As introduced above, adaptive Bayesian methods enable studies to incorporate established data (the “prior”) with new information as data collection progresses. Such an approach can be used to assess the probability of identifying a predefined proportion of patients with abnormal CMi nociceptor activity at the end of recruitment.

In the Bayesian approach, the prior knowledge is represented as a Beta distribution. For example, using data from Evdokimov et al. (2019), where 11 FMS patients had spontaneous activity in CMi fibres and 16 did not, one could define a prior as Beta(11,16).

Alternatively, a more permissive prior of 20% can be used, which incorporates the lower bound of the 95% confidence interval from Evdokimov et al. (2019), but is still almost three times greater than the highest rates reported in healthy aging. Given the uncertainty, i.e., the range of the confidence interval, this prior can be de-weighted by using a Beta(1,4).

The primary analysis will use the de-weighted prior Beta(1,4). Sensitivity analyses will additionally be conducted using a non-informative prior Beta(1,1) and the literature-based prior Beta(11,16) to assess robustness of posterior estimates and stopping decisions to prior specification.

This choice allows new data to meaningfully update the posterior estimate of the proportion of patients with fibromyalgia having spontaneous activity, even with small sample sizes such as in early planned interim assessments.

The final aspect to consider is the confidence in the estimate of the proportion. This is critical when considering interim analysis for futility, as it effectively captures the risk of stopping the study early, when more that 20% of FMS patients do indeed have ongoing spontaneous activity in their CMi fibres – i.e., the risk of a type 2 error, or a false negative.

To understand the impact of implementing a Bayesian approach, 10,000 simulations were performed using the two alternative priors discussed above i.e., Beta(11,16) and Beta(1,4) both at 90%, 95% and 98% confidence of futility, and with actual population prevalences modelled at:

1. 40%, as per Evdokimov et al., 2019
2. 22%, i.e., just over the meaningful threshold, and 3 times the upper estimate of spontaneous activity in healthy aging,
3. 15%, i.e., below the meaningful threshold and within the confidence bounds of health aging.

As shown in Figure 3 (upper panels), the prior (Beta(11,16)) is not meaningfully updated at any interim analysis point, at any confidence. In contrast, with the de-weighted prior (Beta(1,4)), futility is never detected when the population rate is 40%, and only occurs at <10% of simulations, with the most permissive confidence when the population rate is 22%. Thus it appropriately enables recruitment to proceed when the population rate is at or above our meaningful threshold. In contrast, and as shown in Figure 3 (lower panels), when the population rate is below our meaningful threshold, potentially within the bounds of normal aging, futility is detected, with less stringent confidence levels leading to more trials deemed futile.

**Figure 3.**
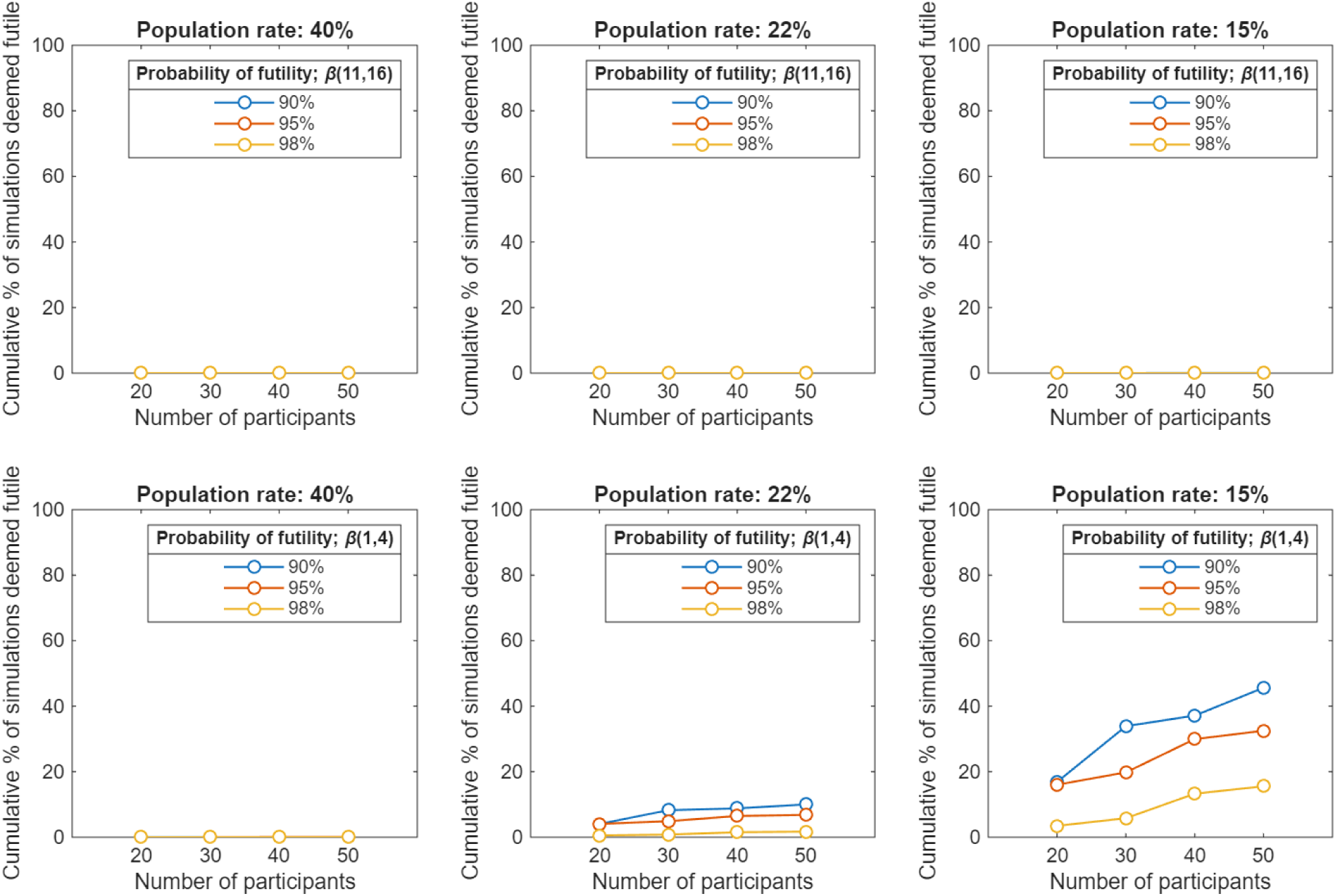
Cumulative percentage of simulations (n = 10,000) where interim assessments applying a Bayesian approach indicated a 90% (blue), 95% (red), or 98% (yellow) probability of futility (where futility is defined as the population rate <20%) and were terminated early when the population rate of spontaneous activity in CMi fibres is 40% (left), 22% (middle), or 15% (right). With the stringent prior (Beta(11,16) shown in the upper three panels and the de-weighted prior (Beta(1,4)) shown in the lower three panels.

A decision on risk vs benefit was therefore required. This was debated within the authorship and with our Public and Patient Involvement and Engagement (PPIE) Group. With the PPIE team, the above simulations, including the priors and confidence intervals, and their impact on the study outcomes were discussed and considered. This group, with lived experience of chronic pain and fibromyalgia, felt:

1. The risks, including side effects and burden of testing, was effectively balanced against the benefits of advancing understanding of pain in fibromyalgia.
2. There is so much uncertainty around the mechanisms of fibromyalgia, and so much uncertainty around how best to treat fibromyalgia, that the risk of futility is worthwhile to add knowledge, even if that knowledge is counter to the hypothesis.
3. If futility could be detected confidently and early, then the trial should stop to minimise risks to patients, and to give them the opportunity to participate in alternate studies.
4. Too stringent a cut off means that we might risk stopping too early, when the abnormalities are present. This was felt to be a particularly undesirable outcome.

Given these simulations, the input of the PPIE team and the consideration of the authors it was decided that *if the probability of futility (i.e., the probability that the prevalence is less than 20%) exceeds 95%, then the study will be deemed futile and the study terminated early*. At 95%, the risk of type 2 error (i.e., incorrect early termination) is minimised, but is sufficiently effective when the population prevalence is low. Interim analyses will be conducted after the first 20 evaluable patients (i.e., patients with at least one classified CMi fibre), and subsequently after every additional 10 evaluable patients. Recruitment will continue until either futility is reached or the funded maximum of 60 patients are recruited.

Only evaluable patients (i.e., those with at least one successfully classified CMi fibre) will contribute to the primary Bayesian updating. Patients who withdraw before completion of microneurography or in whom no classifiable CMi fibre is obtained will be described but excluded from the primary analysis. The number and reasons for exclusion will be reported.

The strategy can be summarised as follows. Let *θ* denote the true prevalence of spontaneous CMi activity in fibromyalgia patients. With prior distribution *θ* ∼ *Beta*(*α*_0_, (*β*_0_), and after observing *x* patients with spontaneous activity out of *n* evaluable patients, the posterior distribution is:

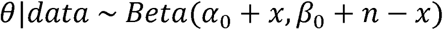

The study will be stopped for futility if:

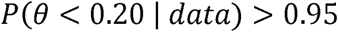

### Secondary analyses

In parallel to the main study, a smaller study in age- and sex-matched healthy controls (n = 25) will be performed to ensure that prevalence of spontaneous CMi fibres using this microneurography protocol is comparable to the existing literature (Evdokimov et al., 2019; Namer et al., 2009; Serra et al., 2014). This population will also be used to assess if the prevalence of spontaneous activity in CMi fibres is higher in the fibromyalgia population than the healthy population using Fisher’s Exact test.

Further analysis will be conducted to stratify the fibromyalgia population based on QST phenotype, and if the prevalence of spontaneous activity in these groups is different. Patients will be categorised into the irritable and non-irritable nociceptor QST phenotypes, as defined in previous studies (Carmland et al., 2024; Demant et al., 2014). In summary, patients with the irritable nociceptor phenotype exhibit normal cold and warm detection thresholds, as well as one or more test demonstrating gain-of-function including dynamic mechanical allodynia, reduced mechanical or pressure pain thresholds, increased mechanical pain sensitivity, or reduced cold or heat pain thresholds.

We will also use QST phenotypes to determine to what extent, if any, that fibromyalgia patients exhibit altered tactile sensitivity (i.e., tuning fork sensitivity, mechanical detection thresholds, ascending vibration detection thresholds (SenseCheQ)) compared with the healthy control population.

We will finally assess whether the QST profiles of fibromyalgia patients with spontaneous activity differ from those without. Specifically, via the adapted short-form McGill Pain Questionnaire, we will assess if patients with spontaneous activity report higher pain scores in the skin of the foot dorsum, perceive pain as originating from the skin, or use different pain descriptors than patients without ongoing activity. Additionally, we will assess if patients with mechanically evoked activity in CMi fibres report higher mechanically-evoked pain scores, both during the QST session and microneurography, and the correlation with brush allodynia.

### Ethics and dissemination

#### Data access statement

Anonymised patient data and associated code used for this manuscript and the study will be available on reasonable request to the corresponding author.

## Discussion

This protocol represents an important step for studies conducting microneurography with patients. Pre-publication of protocols is increasingly seen as being important for ethical and transparent practice in science, particularly with a technique that can be time consuming and demanding for both researcher and participant (*WMA - The World Medical Association-WMA Declaration of Helsinki – Ethical Principles for Medical Research Involving Human Participants*, n.d.). Additionally, the protocol incorporates a Bayesian approach to minimise patient burden, i.e., balancing the value of replication and ongoing exploration of a peripheral nociceptive drive in fibromyalgia, against the impact on patients of a ∼4 hour protocol with a risk of side-effects, albeit mild and self-limiting. The protocol achieves this whilst adequately balancing the risk of type 2 error.

The microneurography protocol seeks to replicate 1) that CMi fibres will have ongoing, non-evoked activity in fibromyalgia patients, and 2) that CMi fibres develop mechanical sensitivity in fibromyalgia patients. The protocol deviates from prior protocols with microneurography in fibromyalgia patients in two important aspects. Firstly, the classification of the CMi fibres is achieved using a) the low frequency ADS protocol which is b) delivered early in the protocol, with the assessment of ongoing activity commencing immediately following classification. This approach aims to maximise the probability of the hypotheses being tested, whilst balancing that against patient comfort, potential for withdrawal, and testing duration. It also enables the primary hypothesis to be assessed prior to a) mechanical stimulation of the receptive field, which can result in electrode movement and thus loss of the ability to electrically excite the unit, and b) prior to the delivery of 2 Hz stimuli that can be less well tolerated than the low frequency protocol and often leads to fatigue/inability of the unit to follow the stimulation frequency that can complicate subsequent classification.

The protocol tests mechanical sensitivity with a 512 mN von Frey filament. This stimulus strength is higher than in some prior reports, where the aim was to differentiate mechanically sensitive from mechanically insensitive C-fibres (Obreja et al., 2010). 512 mN was selected as it falls below threshold of activation of CMi fibres in healthy participants (Obreja et al., 2010; Serra et al., 2004; Weidner et al., 1999), but is sufficiently intense to identify abnormal mechanical sensitivity in CMi fibres (Ribeiro et al., 2025). It is acknowledged that this protocol will not distinguish CMi fibres from the recently described and rarer very-high-threshold C-nociceptors (VHT) (Werland et al., 2021).

The integration of psychophysical measures of nociceptive and non-nociceptive input, i.e., QST, in the same individuals undergoing microneurography, and in an equivalent anatomic location, is a strength of the protocol. This approach is similar to that employed previously (Beisswanger et al., 2022) in patients with small fibre neuropathy and neuropathic pain. Their findings are relevant, as they sought to examine the relationship between microneurography findings in C-nociceptors and patient report; either of ongoing pain or sensory testing. Interestingly, whilst the increased prevalence of abnormal C-nociceptor activity, including ongoing activity in CMi fibres, was clearly present, the relation to QST measures, including the composite irritable nociceptor classification, was less clear. This includes the identification of hyperactive C-fibres in both the irritable nociceptor and non-irritable nociceptor groups. Given the importance placed on mechanism-based stratification and treatment (e.g., (Raja et al., 2020)), and the use of QST to determine these mechanisms (e.g., (Carmland et al., 2024)), it is important to assess the relationship between directly measured nociceptor activity and QST profiling, particularly across different chronic pain aetiologies.

### Comparisons with other designs

The alternative to a Bayesian approach is to use a classical frequentist approach, where the probability of futility is assessed without incorporation of a prior. We can directly contrast the frequentist approach by repeating the simulations shown previously for the Bayesian approach. As can be seen in Figure 4, using the same 95% confidence level as above, more than 96% of simulations failed to correctly identify futility using the frequentist approach. In comparison, the Bayesian approach that this protocol will employ correctly stops before full recruitment in 33.3% of simulations (probability of futility at 95%), therefore achieving increased protection for participants.

**Figure 4.**
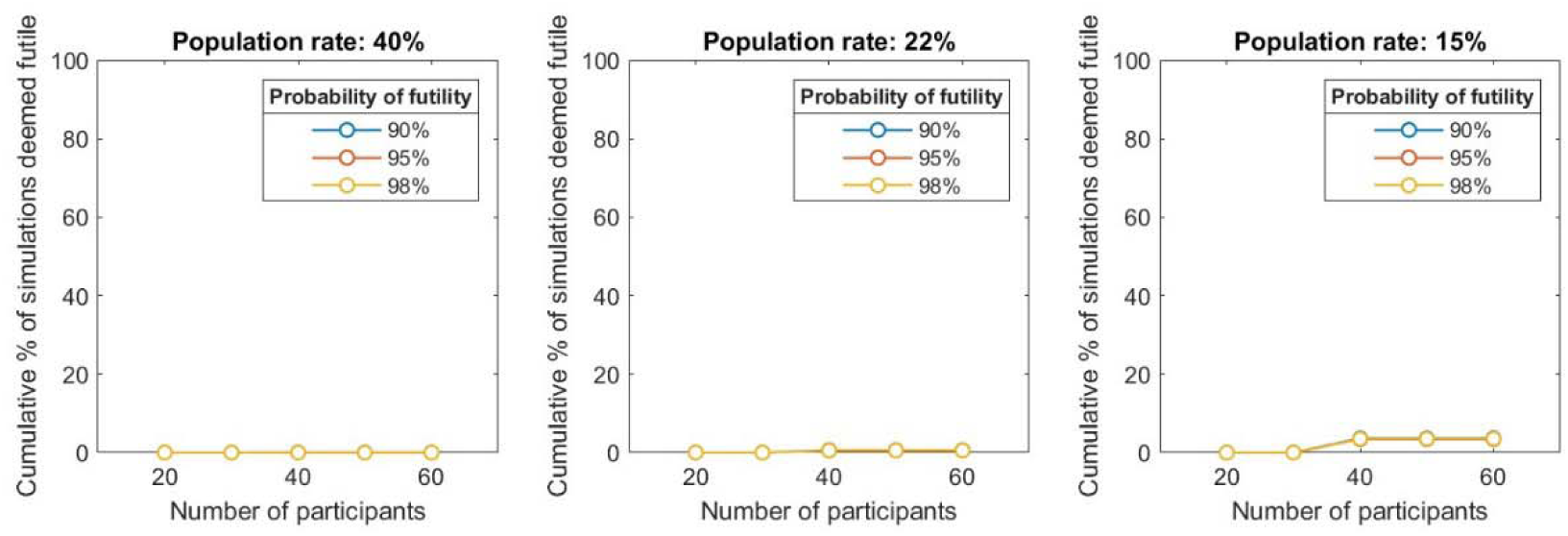
– Percentage of simulations (n = 10,000) where interim assessments applying a frequentist approach (i.e., a normal distribution without prior) indicated 90%, 95%, or 98% probability of futility (where futility is defined as the population rate <20%) when the population rate of spontaneous activity in CMi fibres is 40% (left), 22% (middle), or 15% (right).

Another alternative is to compare the prevalence in the fibromyalgia population and the healthy population. Taking the midpoint for the estimates of the prevalence of spontaneous activity in CMi fibres of ∼5% of healthy controls and the estimate of ∼40% of fibromyalgia patients, a conventional frequentist study design would need to enrol 46 participants (23 in each arm) to achieve sufficient power (α of 0.05 and a confidence of 80%) to demonstrate statistically significant difference (G*Power 3.1, Fisher’s Exact test) (Faul et al., 2007), noting that the results in healthy controls might be unreliable due to the very low prevalence. If the actual proportion of patients and controls with spontaneous activity in CMi fibres are closer than predicted, but still within the confidence intervals cited earlier, then the required sample size to reach power increases substantially. For example, with the prevalence at ∼8% and ∼22% for the healthy control and fibromyalgia populations respectively, a study would require 220 participants (110 in each arm) to reach the same power (G*Power 3.1, Fisher’s Exact test) (Faul et al., 2007). Recruiting such numbers is unlikely to be feasible in a single centre microneurography trial and thus the frequentist approach risks inconclusive outcomes and/or false negatives.

### Statistical considerations

Interim analyses are often employed to test a comparative hypothesis, for example, “*the proportion of abnormal activity in patients is higher than that in controls*”. In such a case, multiple statistical comparisons will be made and adjustments are required to account for this, for example alpha spending (O’Brien & Fleming, 1979). In this protocol, the interim analyses are used to predict the probability of futility, not for statistical comparisons, and so adjustments for multiple comparisons are not required. The outcome that will govern continued recruitment, or stopping for futility, is the probability that the prevalence of spontaneous activity in Type 1B/CMi fibres, at the end of recruitment (n = 60 patients) will be above 20%.

### Concluding remarks

The Bayesian approach to a clinical study described here will allow thorough assessment of the contribution of electrophysiological abnormalities to the pain experience of fibromyalgia patients. Furthermore, this approach will maximise both the study efficiency and impact of the findings whilst minimising the burden on the patient population.

## Authors’ contributions

JPD and AEP contributed to the design and development of the protocol. FK and AB contributed to the interim analysis methods and simulations. EAA performed the simulations. EAA and JPD wrote the manuscript with input from all authors.

